# Engagement with communities at-risk of scrub typhus: lessons learned from Northen Thailand

**DOI:** 10.64898/2026.05.16.26353374

**Authors:** Carlo Perrone, Nipaphan Kanthawang, Sue J Lee, Wanirud Horcharoen, Tripop Phowkanta, Phaik Yeong Cheah

## Abstract

**Background:** In northern Thailand, scrub typhus primarily affects rural and hill tribe communities, yet awareness is low. In 2020 we trained community health volunteers (CHVs) to raise awareness in their communities using a train-the-trainer approach. CHV knowledge rose dramatically. However, we did not assess the effectiveness in community members and our strategy relied heavily on CHVs, who have limited availability.

**Methods:** In a second phase, object of this manuscript and conducted in 2022-2023, after training CHVs we measured the effectiveness in community members and compared in-person sessions carried out by CHVs with sessions using video or audio recordings only. All materials included key information about scrub typhus transmission, symptomatology, prevention, and management and had been developed following feedback from participants from the first round of activities in 2020. Effectiveness was evaluated using a questionnaire assessing scrub typhus knowledge. CHVs were also asked to rate the feasibility of suggested preventative measures.

**Results:** A total of 74 community members participated in six sessions. Knowledge of scrub typhus significantly improved post-training (median score increased from 2 to 6 out of 9, p<0.001) and audio and video recordings were as effective as in-person trainings. CHVs noted that some of the preventative measures recommended are difficult to put in practice such as wearing gloves, washing daily after work, avoiding kneeling and resting on the ground, and washing clothes daily.

**Conclusions:** Our findings support the use of locally adapted multimedia training for scrub typhus prevention, showing that scalable formats can be as effective as in-person ones. Further collaborative work with people at risk should refine preventative messages to improve feasibility and acceptance.

## Background

Scrub typhus, a febrile vector-borne disease, is one of the most common causes of fever in the Asia-Pacific region. People can reduce the risk of acquiring it through preventative behaviours and, if recognized and treated early, life-threatening complications can be avoided.

However, awareness among people at risk is low. Northern Thailand and in particular Chiang Rai, its northernmost province, has a particularly high incidence (Wangrangsimakul, Elliott et al. 2020). Predominantly rural, the risk of disease is especially high for those who farm its hills, whom we identified as the key population to engage with (Wangrangsimakul, Kanthawang et al. 2020). In 2019 we partnered with the primary care department of Chiang Rai Prachanukroh Provincial hospital, and carried out the first phase of our engagement sessions with 117 community health volunteers (CHVs) and 17 primary care unit (PCU) workers from the rural parts of Mueang district (Perrone, Kanthawang et al. 2024). The aim was to engage the healthcare workers and CHVs, so they could in turn provide health education to the wider community. For these activities we created training materials including a video and a flipchart depicting key messages on scrub typhus epidemiology, clinical features, management, and preventative behaviours. Following feedback from the CHVs and PCU workers, an audio recording was developed and the original materials were improved. In interactive sessions, the materials were reviewed and demonstrated to the participants who were then given access to them to use at their discretion within communities. The sessions in 2020 successfully raised knowledge of scrub typhus among the CHVs and PCU workers, with almost all participants achieving the maximum score on the knowledge test at 6 months post-training, irrespective of their baseline score.

While in the first phase in 2020 our evaluation focused on CHVs and PCU workers, in this second phase, we aimed to measure the success of trained CHVs in raising scrub typhus knowledge among community members. The second phase was conducted in a different district of Chiang Rai Province using the same approach. In addition, three different training procedures (each using different materials, see below) were compared with one another and feedback on the feasibility of suggested preventative measures was collected. This manuscript describes the second phase, presents its findings, and contextualises them in relation to the first phase.

## Materials and methods

### Project materials

For this project we used a portable flipchart, a video, and an audio track, intended to aid CHVs in transmitting scrub typhus knowledge to their communities. The materials all focused on scrub typhus and provided consistent messaging on manifestations, transmission, prevention, recognition, and when/where to seek treatment. The flipchart consisted of 14 pictures drawn by a local artist representing the key concepts (e.g. disease symptoms or risk environment) on one side for the audience and a longer explanation on the flip-side for the trainer; the illustrations were made to reflect the real-life context of hill tribe groups.

The video is 6 minutes and 30 seconds long. It tells the same key concepts in a narrative fashion using simple words and shows footage related to the messages (e.g. aerial shots of risk environment, a person donning protective clothing). It was especially shot for this project with non-professional actors, some images from the flipcharts and photos by our research groups were also included. The video is available in the most commonly used languages in this province i.e. Thai, Akha, Lahu, and Karen.

The audio track consists of only the sound-recording from the video, has therefore the same length and is also available in Thai, Akha, Lahu, and Karen. The materials can be accessed on zenodo.com (Wangrangsimakul, Kanthawang et al. 2020, Wangrangsimakul, Kanthawang et al. 2023).

### Project activities

The second phase was carried out between July 2022 and March 2023. It consisted of twelve engagement sessions, five at different PCUs, one at the district hospital; and six in rural villages (see below). All sessions were in Mae Suai district, Chiang Rai Province. Mae Suai is mostly rural, hilly, it borders Myanmar to the west, and its landscapes are rich in coffee plantations and orchards. It was chosen due to the high incidence of scrub typhus and high proportion of hill tribe ethnic groups.

The engagement sessions with CHVs and primary care unit workers were carried out as previously described (Perrone, Kanthawang et al. 2024). Briefly, there was a presentation on scrub typhus and a role-play session where participants simulated training community members with one another (train the trainers), using the flipchart described above. Participants were encouraged to share their knowledge about scrub typhus, using the provided materials if they considered them helpful.

In the second phase, we added a component. At the end of the training, 6 participants (CHVs) volunteered to have the project team accompany them in their villages to observe how they would relay what they had learned. Two of the participants were asked to deliver training using the provided flipchart, two to only use the video, and two to use the audio track. The six CHVs were informed about which materials would be used and were asked to invite community members as participants at their discretion.

Before and immediately after the training provided by these six participants in their villages, community members were asked to complete a questionnaire assessing their knowledge of scrub typhus. The questionnaire consisted of 9 questions on key messages included in the materials (Suppl 1). Community members who were not capable of reading or writing in Thai language were helped by the project team, CHVs, or peers. Key features and differences between the first and second phase are presented in Table 1.

**Table 1.** Description of study design, key characteristics and differences (underlined) between phase one and phase two.

|  | Summary of project design |  |
| --- | --- | --- |
|  | Phase One* | Phase Two |
| Setting and time-period | Mueang Chiang Rai District<br>June-September 2020 | Mae Suai District<br>July 2022- March 2023 |
| Training of community health volunteers (CHVs) and other healthcare workers | 124 CHVs;<br>17 PCU-workers (nurses, nurse aids, public health officers) | 103 CHVs;<br>31 PCU-workers (nurses, public health officers, care-takers) |
| Materials used | - Large Flipchart<br>- Video (Thai, Akha, Lahu)<br>In later stages, based on participant feedback:<br>- Small portable flipcharts<br>- Leaflets | - Large flipchart<br>- Video (Thai, Akha, Lahu, <u>Karen</u>)<br>- <u>Audio</u> (Thai, Akha, Lahu, Karen)<br>- Small portable flipcharts<br>- Leaflets |
| Assessment of baseline scrub typhus knowledge in CHV and PCU-W | Done | Done |
| Assessment of scrub typhus knowledge increase in CHV and PCU-W | <u>Done</u> | Not done |
| Assessment of scrub typhus knowledge increase in community members | Not done | <u>Done</u> (n=74) |
| Comparison of different approaches (Flipchart, audio, and video) in increasing knowledge | Not done | <u>Done</u> (Audio: n=26; Video: n=31; Flipchart: n=17) |

\* Perrone, Kanthawang et al. 2024

Demographic characteristics of the community members from participating villages were summarized descriptively and scores from questionnaires presented as medians with 25^th^ and 75^th^ percentiles. Comparisons between groups were made using the rank-sum test for scores and the chi-squared test for categorical data.

To analyse the effectiveness of the three training procedures in increasing scrub typhus knowledge among community members overall and comparatively, Wilcoxon’s matched pairs signed rank test and linear regression were carried out, respectively. In addition to baseline score, other variables considered for inclusion (age, education, ethnicity) in the adjusted regression were chosen based on findings from the first phase of our training in 2019 (Perrone, Kanthawang et al. 2024). As age was strongly correlated with education levels (with older participants less likely to have received any education) we used Akaike’s information criterion to determine which provided a better fit for the final model. For all tests, a p-value lower than 0.05 was considered statistically significant.

We asked 20 randomly selected CHVs from the engagement sessions to rank the feasibility of the preventative measures that were proposed in the training materials by grading each of them from 1 (very easy) to 5 (very difficult). These questions were asked by telephone and compiled by the project team.

## Results

### Effectiveness of the materials in raising scrub typhus knowledge in community members

Between February and March 2023, we observed 6 training sessions held by CHVs with their communities. Two of the sessions used oral presentations aided by flipcharts, two used the audio track, and two used the video. A total of 74 community members attended, ranging between 7 and 19 community members in each session. The median age was 52 years, 51 (69%) community members were female, and 41 (53%) were non-Thai (Table 2).

**Table 2.** Demographic characteristics and scrub typhus knowledge scores by training method.

| Characteristic | Training Method |  |  | Total, n (%) |
| --- | --- | --- | --- | --- |
|  | Flipchart,<br>n (%) | Video,<br>n (%) | Audio,<br>n (%) |  |
| Total | 17 | 31 | 26 | 74 |
| Gender |  |  |  | *p=0.367* |
| - Female | 14 (82) | 19 (61) | 18 (69) | 51 (69) |
| - Male | 3 (18) | 12 (39) | 7 (27) | 22 (30) |
| - Missing | 0 | 0 | 1 (4) | 1 (1) |
| Age |  |  |  | *p=0.001* |
| - <30 | 1 (6) | 14 (45) | 2 (8) | 17 (23) |
| - 30-49 | 5 (29) | 8 (26) | 5 (19) | 18 (24) |
| - 50-64 | 8 (47) | 5 (16) | 9 (35) | 22 (30) |
| - ≥65 | 3 (18) | 4 (13) | 10 (38) | 17 (23) |
| Formal education (primary school or higher) |  |  |  | *p= 0.015* |
| - Yes | 14 (82) | 18 (58) | 8 (32) | 40 (55) |
| - No | 3 (18) | 13 (42) | 17 (68) | 33 (44) |
| - Missing | 0 | 0 | 1(4) | 1 (1) |
| Ethnicity |  |  |  | *p<0.001* |
| - Thai | 17 (100%) | 1 (3%) | 17 (65%) | 35 (47%) |
| - Akha | 0 | 16 (52%) | 3 (12%) | 19 (26%) |
| - Lahu | 0 | 13 (42%) | 6 (23%) | 19 (26%) |
| - Other | 0 | 1 (3%) | 0 | 1 (1%) |
| Knowledge score* |  |  |  |  |
| - Baseline |  |  |  | *p*=0.031 |
| Median (p25, p75) | 1 (0-3) | 1 (0-2) | 2 (2-4) | 2 (1-3) |
| - Post-training |  |  |  | *p*=0.007 |
| Median (p25, p75) | 6 (4-7) | 7 (6-8) | 5 (3-6) | 6 (4-8) |
| Score increase |  |  |  | *p*<0.001 |
| - Mean (SD) | 3.9 (1.9) | 5.2 (1.9) | 2.3 (1.8) | 3.9 (2.2) |

\* Out of a maximum score of 9 SD: Standard deviation

Overall, the flipchart-aided sessions were attended by the smallest number of community members (n=17) whereas nearly twice as many (n=31) attended the ones using the video. Also, in the flipchart group, all participants were Thai. Those who attended the video training were younger than those who were trained using the other two methods (Table 2). The proportion of community members without formal education was highest in the Audio group.

Community members had a very low baseline knowledge of scrub typhus, with an overall median score of 2/9 (p25-p75 1-3) on the questionnaire. The maximum score was 7/9 (n=2 community members) and 16/74 (21.6%) community members scored 0/9. After the training sessions, the median scored increased to 6/9 (p25-p75 4-8, p<0.001 paired comparison with baseline score). The average increase in knowledge overall was 3.9 points, with three people scoring 100% (9/9). Median scores before and after the sessions for the three different methods are represented in Figure 1. Linear regression adjusted for the baseline score showed that participants in the video training session scored on average 1.21 (CI 0.24-2.18, p=0.015) points higher on the post-training questionnaire when compared to training using flipcharts (Table 3).

**Table 3.** Logistic regression of post-training score.

|  | n | Univariate regression coefficient<sup>a</sup> | p-value | Multivariable regression coeff.<sup>b</sup> | p-value |
| --- | --- | --- | --- | --- | --- |
| Training material |  |  |  |  |  |
| - Flipchart/ frontal | 17 | Ref. | - | Ref. | - |
| - Audio | 26 | -0.99 (-2.02; 0.034) | 0.058 | -0.78 (-1.89; 0.33) | 0.166 |
| - Video | 31 | 1.21 (0.24-2.18) | 0.015 | 0.36 (-1.19; 1.92) | 0.642 |
| Ethnicity |  |  |  |  |  |
| - Thai | 35 | Ref. | - | Ref. | - |
| - Akha | 19 | 1.43 (0.46; 2.39) | 0.004 | -0.17 (-1.80; -1.46) | 0.836 |
| - Lahu | 19 | 1.44 (0.42; 2.45) | 0.006 | 0.62 (-0.81; 2.04) | 0.392 |
| - Other | 1 | 4.44 (1.03; 7.85) | 0.012 | 2.42 (-1.10; 5.94) | 0.174 |
| Age | 74 | -.05 (-0.07; -0.03)<sup>c</sup> | <0.001 | -0.35 (-0.64; -0.01)<sup>b</sup> | 0.021 |
| Formal education |  |  |  |  |  |
| - No | 33 | Ref. |  |  | - |
| - Yes | 40 | 1.0 (0.1; 1.8) | 0.021 | - |  |

a: Adjusted for pre-training score b: Adjusted for pre-training score, ethnicity and age c: Per year of age increase

**Figure 1.**
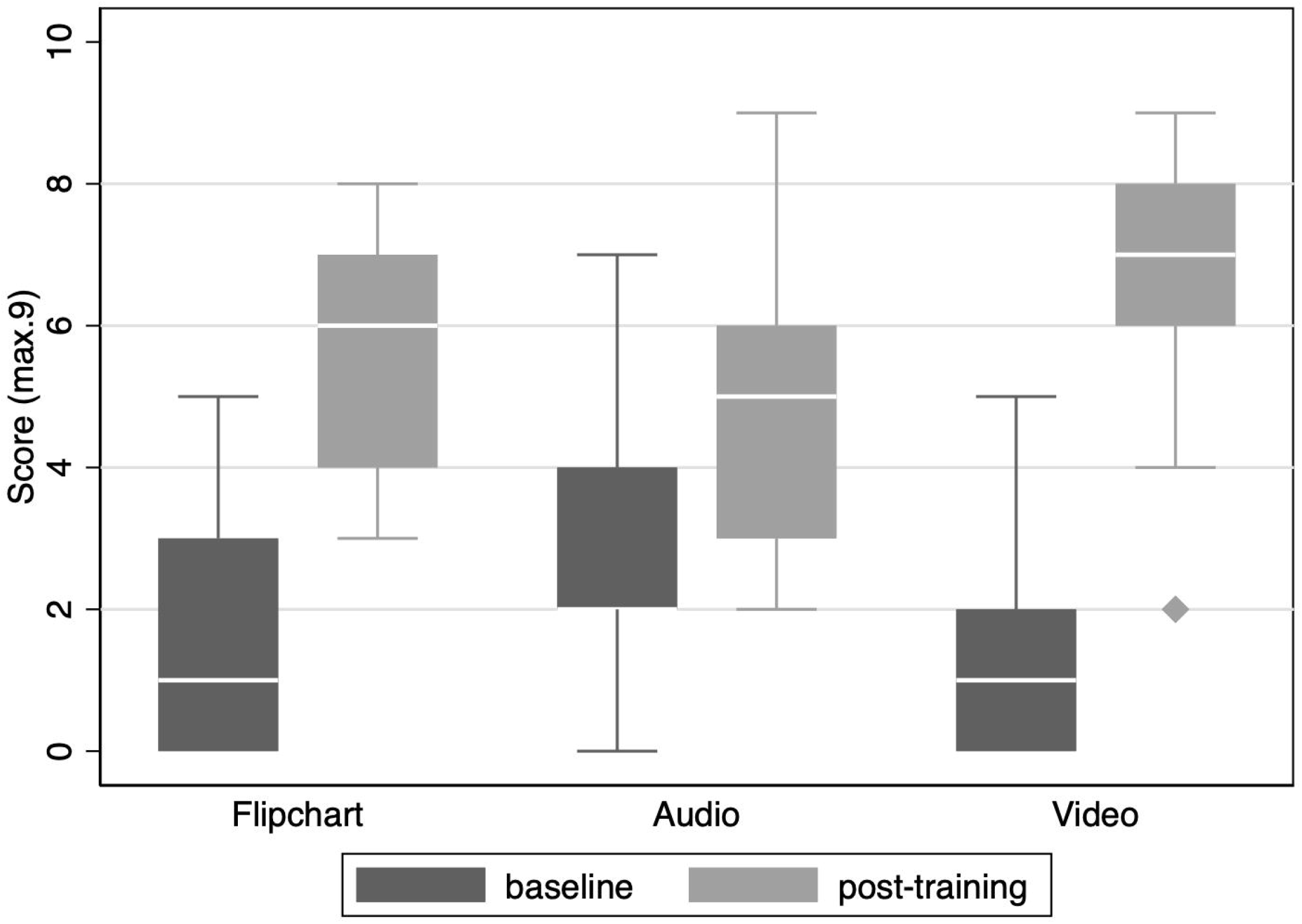
Box and whiskers plot of scrub typhus knowledge scores before and after the training, by training method

People of non-Thai ethnicity were also found to score higher, but in the multivariable model the score difference among training methods and ethnicities was not statistically significant, and age was the only variable associated with a change in post-training score (p=0.021), where younger participants were more likely to score higher (Table 3).

### Perceived feasibility of the proposed preventative measures

In the materials provided for the training sessions, the following were proposed as effective scrub typhus prevention techniques during or after exposure to risk environments: Wearing long sleeved shirts, long trousers, socks and boots, scarves and hats, bathing or showering thoroughly at the end of the day, not kneeling or laying on the naked earth, washing work clothes at the end of each day. The recommendations were selected based on a review of the literature and known vector behaviour. When the 20 randomly selected CHVs were asked about the feasibility of each of the proposed measures, the majority considered wearing protective clothing while working feasible, with the exception of gloves. However, washing daily after work, avoiding kneeling and resting on the ground, and washing clothes daily was regarded as difficult and not practical, especially the latter where nearly 70% ranked daily clothes washing as somewhat or very difficult. These findings are presented in Figure 2.

**Figure 2.**
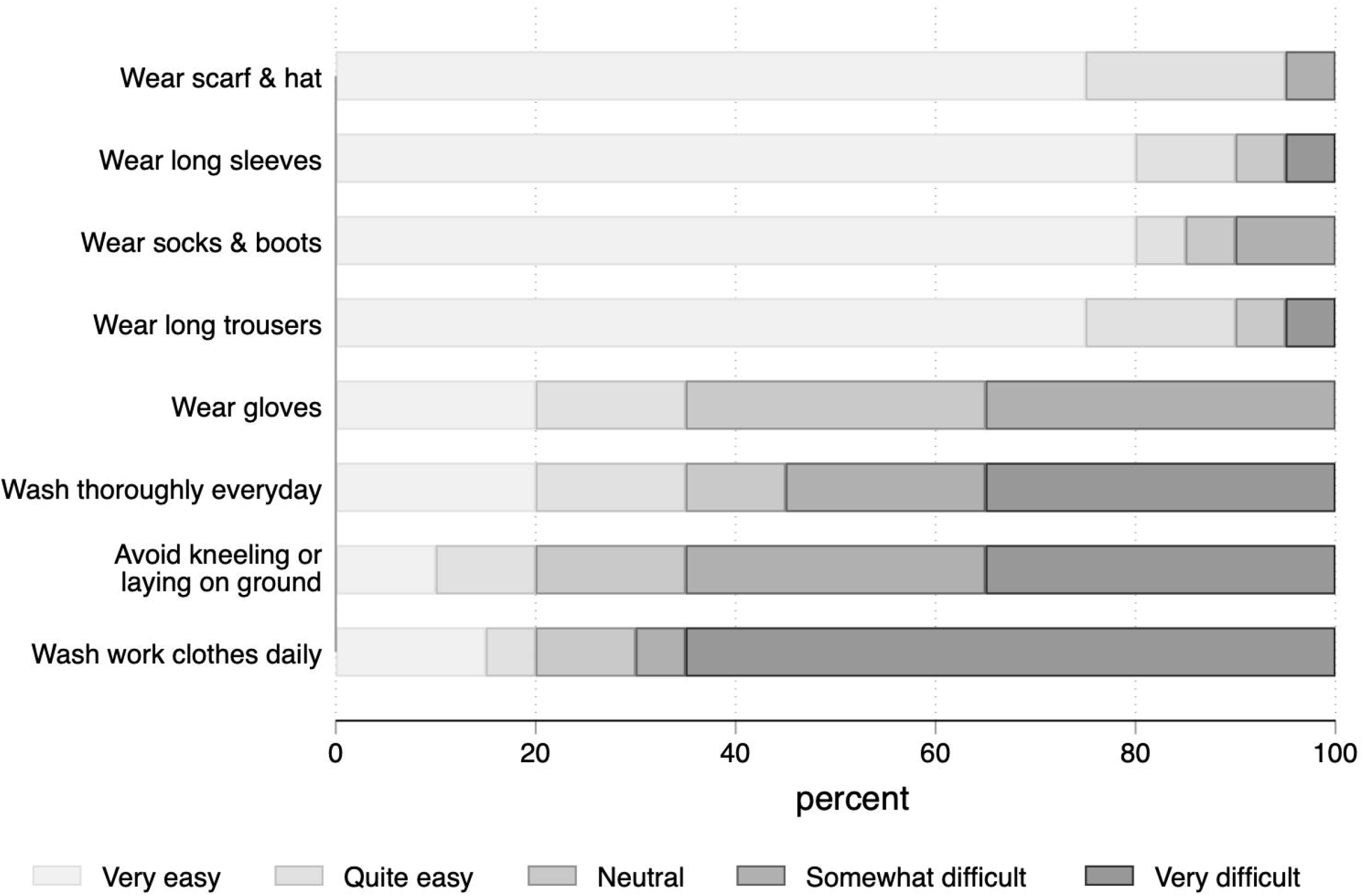
Feasibility of preventative measures as judged by community health volunteers

## Discussion

Overall, the trainings by community health care volunteers improved scrub typhus knowledge among community members. When comparing the three methods, the video appeared to be more effective than both the audio recording or the flipchart. This difference, however, was not significant after adjusting for age and ethnicity. The increase in score among people of non-Thai ethnicity was not significantly different to that of Thais, suggesting that the materials developed in hill tribe languages were at least as effective as the ones in Thai. However, the flipchart training group comprised only Thai people, so ethnicity-related conclusions on this group cannot be made. Importantly, the trainings carried out by community health volunteers using the flipcharts were not superior to any of the other two methods, which suggests that relaying information through video screenings, radios, or village loudspeakers (a widespread means of communication in rural southeast Asia) could be an effective and easily up-scalable strategy.

Notably, none of the trainings with community members in this second phase (2022) achieved the same success as the ones carried out with CHVs in the first phase (2019), despite using the same materials. The main differences between the two stages (the project team training CHVs versus CHVs training community members) were that when CHVs were trained: 1-They had to practice with one another, and 2-They were asked not only to view but also to critically appraise the materials. Community members viewed the presentations and materials in a passive fashion. It is possible that the higher level of participation that was intrinsic to the training of CHVs and the repetition of the messages (by viewing the materials and listening to fellow CHVs practice several times) contributed to the higher success.

Unfortunately, our findings show that some of the preventative measures we are recommending could be hard to put in place, as judged by CHVs. Gloves, for instance, were considered bothersome when picking produce such as coffee or fruits and there being no dedicated places to rest in fields or natural settings, laying directly on the ground is difficult to avoid. Similarly, washing thoroughly one’s body and work clothes daily after a long day of work, often, especially during the rainy and cold season was considered difficult. These behaviours were found (albeit inconsistently) to affect the risk of scrub typhus(Kim, Kim et al. 2008, Tasak, Apidechkul et al. 2023). Focus group discussions with community members could be a useful tool to tailor recommendations to the local context or to find reasonable practicable alternatives and to make sure measures are respectful of context and lifestyle, as has been advocated to overcome barriers to malaria prevention (Naserrudin, Lin et al. 2023).

One limitation of the project was that CHVs were free to invite community members as they saw fit. We chose this approach to reflect how CHVs normally work but due to selection bias generalisability may be reduced. Another limitation is that the audio track was played to a concentrated sitting audience rather than through village public announcement (PA) systems to people going about their regular business, which is how health messages are normally conveyed to communities. Messages through PA systems are often played repeatedly, so the overall effectiveness is likely to be similar, if not higher than what is reported here.

## Conclusions

Our experience supports the use of materials created through community engagement to increase scrub typhus knowledge and shows that audio and video recordings can be at least as effective as in-person trainings with the flipcharts. These methods represent an inexpensive and easily up-scalable resource for rural, low-income communities. Use of video appeared to be superior to other methods, in particular for the younger generation. When participation in training was interactive and messages were repeated several times, the increase in knowledge was more pronounced.

Future efforts should also be directed to discussing preventive behaviours together with people at risk to devise recommendations as feasible as possible.

## Supporting information

Supplement one - Scrub typhus knowledge questionnaire

## Data Availability

All data produced in the present study are available upon reasonable request to the MORU data access committee

## Declarations

## Abbreviations

CHV: Community Health Volunteer
CI: Confidence interval
PA: Public announcement
SD: Standard deviation

## Ethics approval and consent to participate

The Chiang Rai Provincial Health Office Ethics Committee waived ethical board review as the project was considered low-risk for participants. All participants provided verbal informed consent to participate in the sessions and agreed on having the results published with the information de-identified. The study adhered to the Declaration of Helsinki.

### Consent for publication

Not applicable

### Availability of Data and Materials

The datasets used and/or analysed during the current study are available from the corresponding author on reasonable request.

### Funding

This work was supported by the Wellcome Trust [Grant number 220211/Z/20/Z]. For the purpose of open access, the author has applied a CC BY public copyright licence to any Author Accepted Manuscript version arising from this submission.

### Authors’ contributions

CP, NK, and PYC contributed to the conception and design of the paper; WH, TP, CP, and NK were responsible for the acquisition of data; SJL and CP carried out data analysis; CP, SJL, NK, and PYC interpreted the data; CP drafted the first draft of the manuscript which was revised and approved by all other authors.

## Acknowledgements

Thanks to all participants, and in particular to the staff at the primary care units of Doi Chang, Huai Nam Khun, Pa Daet, Tha Kho, and Tung Prao, and the villages of Baan Pang, Huai Masang, Pa Daeng Lisu, Huai Chompoo and Pang Nam Moon for helping us to carry out the activities.

