## Supplement one - Scrub typhus knowledge questionnaire for "Engagement with communities at-risk of scrub typhus: lessons learned from Northen Thailand"

Supplement one: Scrub Typhus Knowledge Questionnaire for community members

1. How is scrub typhus transmitted to humans?
1. The bite of an animal (cat/dog/mouse)	2. By air (cough, sneezing)
3. The bite of a small insect	4. By eating undercooked food
5. I don’t know/not sure
2. What habitat is NOT associated with scrub typhus?

1. Schools	2. Forest

3. Coffee/tea plantations	4. Rice field

5.I don’t know/not sure

3. What are typical symptoms of scrub typhus?
1. Itching, swollen eyes, breathing difficulties
2. Fever, headache, abdominal pain, cough
3. Watery diarrhea and vomiting many times a day
4. High blood sugar, increased urination, reduced consciousness
5. All of the above
6. I don’t know/not sure
4. Which medicine used to treat scrub typhus?

1. Antibiotics	2. Paracetamol

3. Anti-diarrhoeals	4. Anti-emetics

5.I don’t know/not sure

5. Which of the following occupations has the highest risk for scrub typhus?

1. Police	2. Teacher
3. Doctor	4. Farmer

5.I don’t know/not sure

6. How can you prevent catching scrub typhus?

1. Lie down to rest on the ground at break time

2. Vaccination

3. Wear protective clothing

4. Exercising and eating healthy foods.

5.I don’t know/not sure

7. Which of the following is not recommended?

1. Take a shower soon after returning home from agricultural work.

2. Wear fully-covered clothing, long-sleeved shirts and long pants, even on hot days.

3. Buy medicine at the pharmacy when you feel unwell and think you have scrub typhus.

4. Clean and maintain waste areas and areas around the house regularly.

5.I don’t know/not sure

8. Which of the following is correct regarding management of scrub typhus?

1. If you have any symptoms that suggest scrub typhus e.g. fever and eschar after working in the field, plantation or forest, seek medical help.

2. Ignore symptoms of scrub typhus as it rarely needs treatment.

3. Take medication everyday to prevent scrub typhus.

4. I can use herbal or traditional medicine to treat scrub typhus.

5.I don’t know/not sure

9. Which picture shows the skin lesion (eschar) associated with scrub typhus infection?

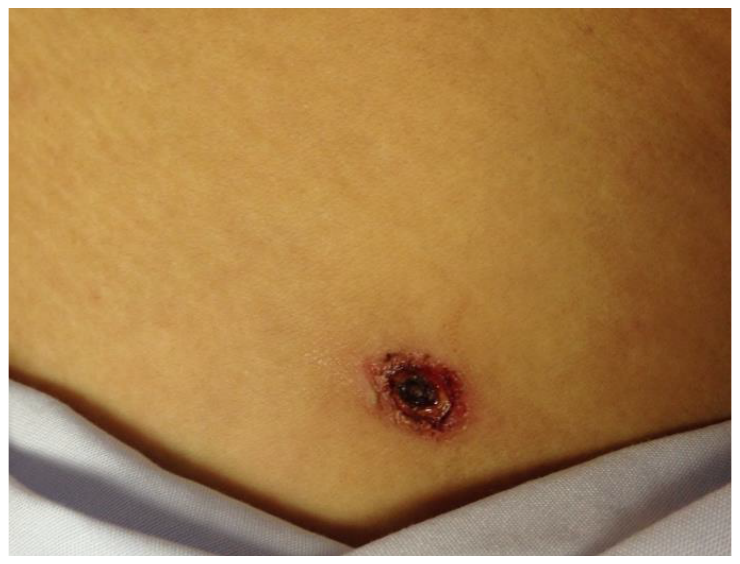

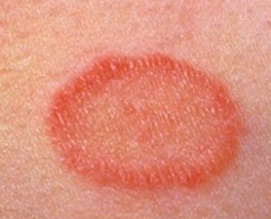
**1).**	2).

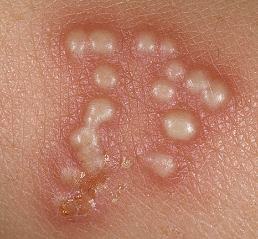

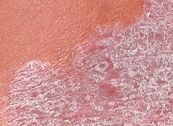
3).	4).
